# Predicting the evolution and control of COVID-19 pandemic in Portugal

**DOI:** 10.1101/2020.03.28.20046250

**Authors:** Ricardo J. Pais, Nuno Taveira

## Abstract

The Coronavirus (COVID-19) is a world pandemic that has been affecting Portugal since 2 of March 2020. Portuguese government has been making efforts to contradict the exponential growth through social isolation measures. We have developed a mathematical model to predict the impact of such measures in the number of infected cases and peak of infection. We estimate the peak to be around 2 million infected cases by the beginning of May if no additional measures are taken. The model shows that current measures effectively isolated 25-30% of the population, contributing for some reduction on the infection peak. Importantly, our simulations show that the infection burden can be further reduced with higher isolation degree, providing information for a second intervention.

## Background

The Coronavirus Disease 2019 (COVID-19) is already considered a world pandemic which is starting to have dramatic effects in Europe where as of 27 of March 265,421 cases have been reported (https://www.ecdc.europa.eu/en/cases-2019-ncov-eueea) [1]. COVID-19 infection in Portugal grows exponentially with an average rate of 34 ± 13 % new cases per day from 2 of March and is far from reaching the peak by the end of March. As of March 27, 4268 infection cases and 76 deaths have been reported (https://covid19.min-saude.pt/wp-content/uploads/2020/03/i026082.pdf). The highest infection burden is found in Porto (317 cases, 7.4%) and in Lisbon (284 cases, 6.7%) but the disease is present in the entire country. As in other countries, infection occurs mostly in individuals’ with ≥40 years of age (71.9% males; 69.3% females). Death cases occur mostly in males (64.5%) all with ≥50 years of age. Predictive models estimate that the peak of COVID-19 infection globally will be between mid-April and May with an estimated total affected of 48 million people [2]. As with most other countries, Portuguese national health care system cannot deal with the increasing demand of care due to limited ventilators and care units [2]. Therefore, the Portuguese government together with the National Health Directorate (DGS) has declared in 18 of March the state of emergency and adopted interventive populational measures (IM) in an attempt to drop the peak of infections even if at the cost of prolonging the infection time. These measures are based on the isolation of people at home-social distancing and adopting protective antiseptic policies. Most forecasting models are based on the number of cases reported and do not take into account the effects of these government-imposed measures and behavioural change. Thus, how these measures impact the evolution of the COVID-19 infection and can prevent the expansion of the epidemic is unknown. Recently published mathematical modelling studies of COVID-19 transmission already provided useful insights that can be used to guide public health measures and resource allocation to better control this pandemic [3,4]. However, most parameters of statistical models have been estimated with high degree of uncertainty, resulting in predictions with wide intervals of confidence [3]. Compartmental models such as SIR models (**S**usceptible, **I**nfected and **R**esistant) are deterministic approaches that have been successful in describing the dynamics of virus infection in populations, including COVID-19 [4,5]. Here, we provide a new SIR model that describe the dynamics of transition of COVID-19 in Portugal during the first 21 days and predicts the impact of the corrective measures towards the expected peak of infection.

## Methods

Basic transmission dynamics of COVID-19 was modelled using a simple mathematical model based on a system of two ordinary differential equations (ODE) developed specifically for this purpose (see equations on Figure 1A). The equations reflect the number of people infected (I) and susceptible (S) to infection per unit of time (dI/dt and dS/dt). In this model, we accounted for the reported average time of duration of infection (τ) of 14 days [6]. Model was calibrated by adjusting the rate constant (k) to approximate the total infection value reported by the DGS at 17 March. No further fitting was performed in this model. The effect of isolating different fractions of the population was modelled through the variation of parameter α in equations. We assumed that protective measures were 99% effective, accounted through model parameter β. The ODEs were solved using PLAS software and series of simulations were carried scanning various values of the α parameter [7]. Simulations were carried with the initial 2 cases reported by the DGS and considering only the population of the grand Lisbon and Porto areas since they represent most of the susceptible population (total of 6.5 ⨯ 10^6^).

**Figure 1.**
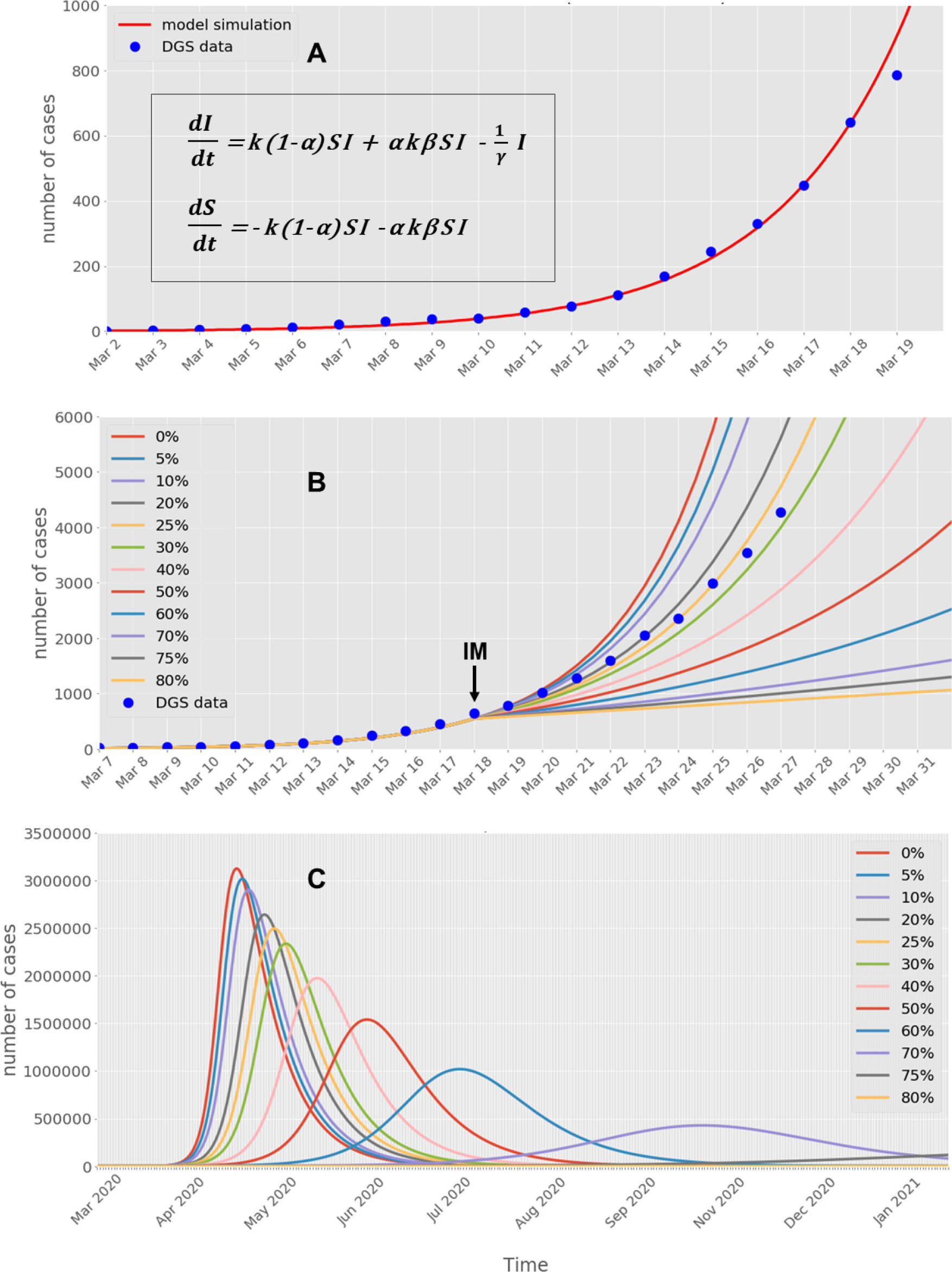
Simulation of multiple scenarios for the dynamics of COVID-19 spreading on Portuguese population. **A)** Model equations and validation with DGS data before applying interventive measures. **B)** Predicted total infected population in the first 21 days for different percentages of isolated population. **C)** Predicted peak of infection for different percentages of isolated population. The IM indicates interventive populational measures. The arrow indicates the time where a change on the model α parameter occur to mimic changes in populations exposed to infection.

## Results

Simulation of the first 18 days with our model was able to describe the exponential increase of the number of confirmed cases reported by the DGS between 2 and 18 of March 2020 (Figure 1A). The predicted peak time for this scenario was 49 days which would be by the 21 of April. This is within the estimated range predicted by statistical modelling of US, Italy and Korea scenarios [2]. Further, the predicted numbers of cases for the end of March if no measures were taken would be around 42 million. This is also in agreement with the number released by the DGS to the social media based on statistical modelling. Thus, the model presented here is consistent with the forecasting made by conventional models, reinforcing the confidence on our model capacity to generate predictions.

Importantly, our results show that the isolation measures had an immediate impact on diminishing the exponential increase of the number of infected cases and this depends on the percentage of the population that is isolated (Figure 1B). This is evident by the increasing deviation of the reported number of cases relative to the unperturbed simulation (0%) with time. The evolution of the number of cases reported by DGS between 18 and 25 of March fit between the simulation curves corresponding to 20% and 30% population isolation. This suggests that the estimated percentage of the population that have been effectively isolated is between these percentages. Interestingly, the data also shows a slight shift with time form being close to the 20% towards 30% suggesting that isolation behaviour of the population was gradual. From simulations, we identify other intervals (e.g. 50-60% and 70-75%) that suggest further isolation percentages may be more effective and still withing a plausible of pandemic time. Based on the fraction of hospitalized and mortality reported by the DGS on 27 March together with our model predictions, we computed several infection indicators for these intervals (Table 1). Model analysis indicates that current government-mandated measures may shift the expected peak of infections towards the beginning of May and can cause a substantial reduction in the infection numbers (Figure 1C, Table 1). Thus, the predicted peak in the number of cases without any isolation measures would be around 2-2.5 million, whereas the intervention measures have decreased it to an estimated 1.2-0.5 million (Table 1). In addition, the estimated reduction of hospitalized patients and death cases on peak would be predicted around 59,000 and 12,000 people, respectively. Our simulations also indicate that the peak of infection can be further reduced ∼3.5-fold with a delay to November if 70% percentage of the population is isolated at home and follows the government recommendations. For higher percentages of isolation (>75%), our model predicts a substantial reduction in the number of infections and delay of peaks, stopping the COVID-19 epidemic. These solutions, would result in much less total mortality and hospitalization requirements on peak in comparison to the current trend (Table 1, Figure 1B-C). Meanwhile, this comes with the burden of prolonging the time of pandemic to almost a year, which can be economically unbearable. In alternative, further isolation to 50-60% of the population may be also a solution that substantially reduce most pandemic indicators and shifts the ending of the pandemic to September, with the peak between June and July.

**Table 1.** Predicted upper and lower values for several COVID-19 infection indicators.

| **Predictions** | **Current control**<br>**(25-30 % isolation)** | **Mild control**<br>**(50-60 % isolation)** | **Optimal control**<br>**(70-75% isolation)** |
| --- | --- | --- | --- |
| **Total infected** | 4,648,087 – 4,791,783 | 3,295,201 – 3,910,457 | 1,354,146 – 2,202,358 |
| **Total mortality** | 41,594 – 44,421 | 18,141 – 27,406 | 2,723 – 7,623 |
| **Infected**<br>**(on peak)** | 2,335,835 – 2,494,627 | 1,018,771 – 1,539,093 | 152,938 – 428,124 |
| **Hospitalized**<br>**(on peak)** | 193,740 – 206,911 | 84,499 – 127,656 | 12,685 – 35,509 |
| **Expected**<br>**Peak occurrence** | 1–10 May 2020 | 6 Jun – 8 Jul 2020 | Oct 2020 – Jan 2021 |

Although our model was precise on describing the exponential curve and explains the shift in the temporal evolution of DGS data, it has limitations that may compromise the exact values of predictions. The fact that we only assume two compartments (Susceptible and Infected) considering the main populated cities (Lisbon and Porto) as one is huge approximation that neglects regional dynamics. Thus, the model is just an approximation that reflects an average trend and may fail to explain regional observations. In this model we also neglected many important parameters of infection transmission such as age groups, social interactions, contact dependent probability, and viral load dependent probability [8]. The inclusion of these parameters would definitely make the model more realistic. However, this data is not available for the Portuguese case and these models require accurate processing of data curation for suitable validation. We have bypassed these limitations by aggregating all of these parameters into one constant, which was fitted to the available data. Overall, the predictions shown here should be taken as semi-quantitative estimates within an upper and lower case-scenarios.

## Conclusions

In this work we demonstrate the potential of modelling COVID-19 dynamics of infection as a useful support tool for predicting the impact of corrective measures. Government-mandated measures to isolate the Portuguese population at home effectively prevented COVID-19 from reaching dramatic numbers in Portugal but still can be substantially improved to reduce the infection peak Our estimates may help guiding additional measures to control the COVID-19 epidemic in Portugal.

## Data Availability

All data is available on request.

